# Extending A Chronological and Geographical Analysis of Personal Reports of COVID-19 on Twitter to England, UK

**DOI:** 10.1101/2020.05.05.20083436

**Authors:** S Golder, Ari Z. Klein, Arjun Magge, Karen O’Connor, Haitao Cai, Davy Weissenbacher, Graciela Gonzalez-Hernandez

## Abstract

The rapidly evolving COVID-19 pandemic presents challenges for actively monitoring its transmission. In this study, we extend a social media mining approach used in the US to automatically identify personal reports of COVID-19 on Twitter in England, UK. The findings indicate that natural language processing and machine learning framework could help provide an early indication of the chronological and geographical distribution of COVID-19 in England.

## Introduction

Given the uncertainty about the trends and extent of the rapidly evolving COVID-19 outbreak, and the delay in much promised testing in the United Kingdom, our understanding of the spread of COVID-19 is limited. The lab-confirmed cases that are released daily are those of only hospital patients with a medical need and more recently healthcare workers (https://coronavirus.data.gov.uk/). An effort for an alternative source of data has emerged in the form of an app – ‘COVID 19 Symptom Tracker’, designed around passively collecting health data on a daily basis from its users (https://covid.ioinzoe.com/data). We propose a different, potentially complementary approach to actively collect reports of COVID=19 posted on Twitter, automatically assessing probable and possible cases by region. As of January 2020 there were 16.7 million Twitter users in the UK (https://www.statista.com/statistics/242606/number-of-active-twitter-users-in-selected-countries/), and using such a platform and free-text reports could facilitate widespread collection of near real-time reports. Our preliminary analysis shows that Twitter data showed an increase in cases 7–14 days ahead of the official reports. Whilst Twitter users may not be posting to aid symptom monitoring, users often describe symptoms or whether they have or suspect they have COVID-19. Using Twitter may also capture reports from a different demographic, people reluctant to use an app, and provide complementary data to official reports and data from the symptom tracker app. This social media mining approach has already been applied to Tweets from users in the United States to analyze the daily trends in the potential exposure to COVID-19 in different states in the US over time^2^.

We proposed to test the feasibility of this methodology on Tweets with a geographical location of England in the UK. To our knowledge, this is the first use of real-time personal reports in Twitter data from England to track COVID-19.

## Methods

The study on the United States used natural language processing and machine learning framework to advance the use of Twitter data as a complementary resource “to understand and model the transmission and trajectory of COVID-19”^1^. We proposed that this methodology could not just be used in the United States but worldwide, and thus begun our case study with England, UK.

The methods to collect tweets (excluding retweets) from the Twitter Streaming API that indicate that the user or a member of the user’s household had been exposed to COVID-19 are the same as used in the U.S. study but the tweets were required to be geo-tagged or have profile location metadata in England in the UK^2^.

We manually annotated a random sample of tweets collected between the 23^rd^ of January 23, 2020 and the 6^th^ of April 2020. Annotation guidelines were developed to help two annotators distinguish tweets that indicate one of three distinguishing categories;

1. Probable: The tweet indicates that the user or a member of the user’s household has been diagnosed with, tested for, denied testing for, symptomatic of, or directly exposed to confirmed or presumptive cases of COVID-19.
2. Possible: The tweet indicates that the user or a member of the user’s household has had experiences that pose a higher risk of exposure to COVID-19 (e.g., recent traveling) or exhibits symptoms that may be, but are less commonly, associated with COVID-19.
3. Other: The tweet is related to COVID-19 and may discuss topics such as testing, symptoms, traveling, or social distancing, but it does not indicate that the user or a member of the user’s household may be infected. Inter-annotator agreement was high^2^. We analysed geographical and temporal trends of posts in category 1 or 2.

## Results

Whilst some posts could only be ascertained as from England, the majority had more specific geographical locations, such as city, county or region. In order to compare the data to that of UK government figures we coded each city to its appropriate region, for instance, Leeds or West Yorkshire were placed in the ‘Yorkshire and Humber’ region. Around 90% of the posts merely discuss COVID-19 (category 3), however, 2065 with a location of England were a probable or possible case of COVID-19 (category 1 or 2).

Figure 1 illustrates the number of detected users from each region in England who have posted “probable” or “possible” tweets between January 23, 2020 and April 6, 2020 and compares this to the UK government statistics for confirmed cases by region in England^3^. Figure 2 illustrates the number of cumulative users in England by report date in conjunction with UK government figures for cumulative cases and cumulative deaths^3^. Figure 3 illustrates the cumulative number of users posting “probable” or “possible” tweets from London compared to other regions in England by report date, January 23, 2020 to April 6, 2020.

**Figure 1.**
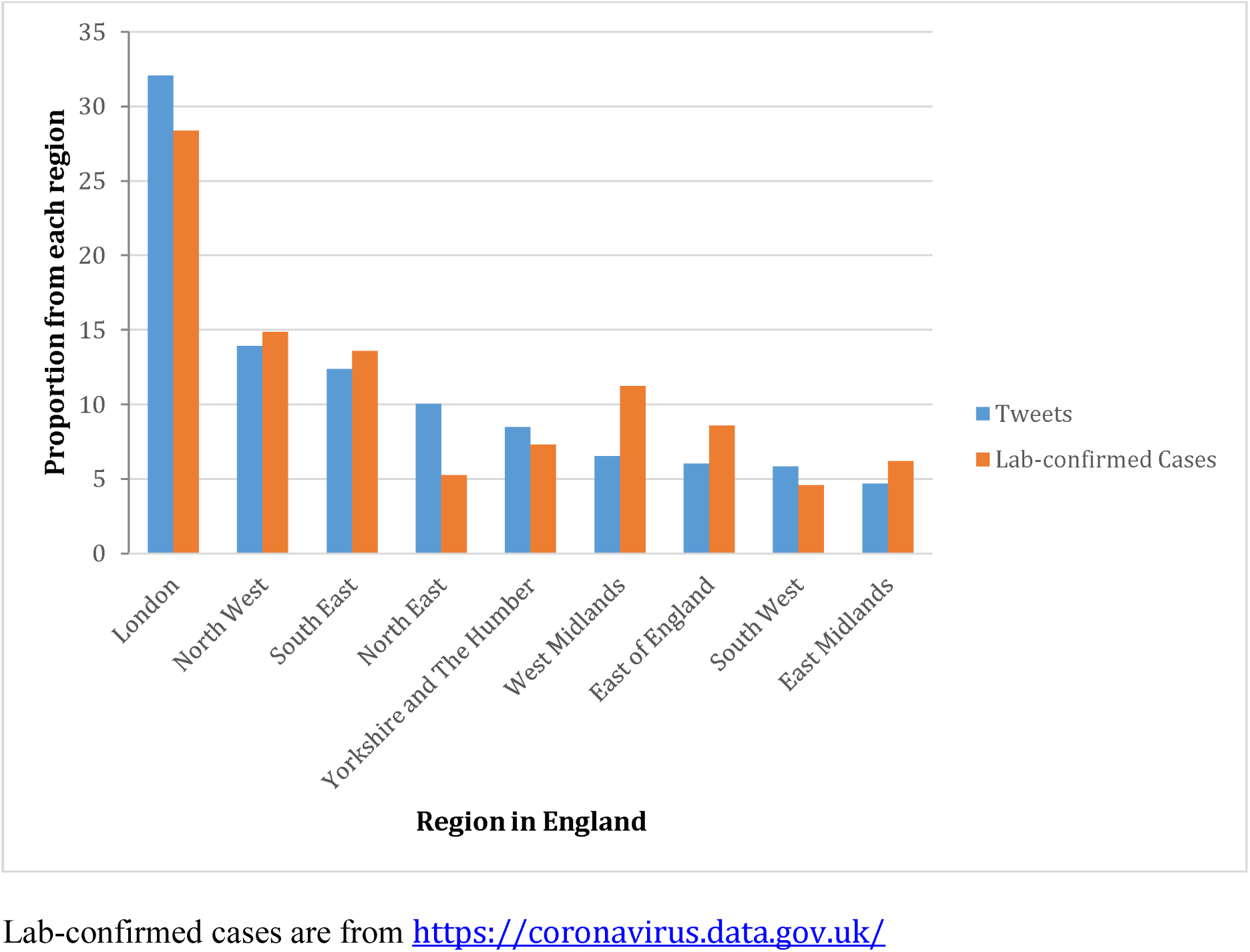
Proportion of all users posting “probable” or “possible” tweets by region, compared to proportion of all government confirmed cases by region, January 23, 2020 to April 6, 2020

**Figure 2.**
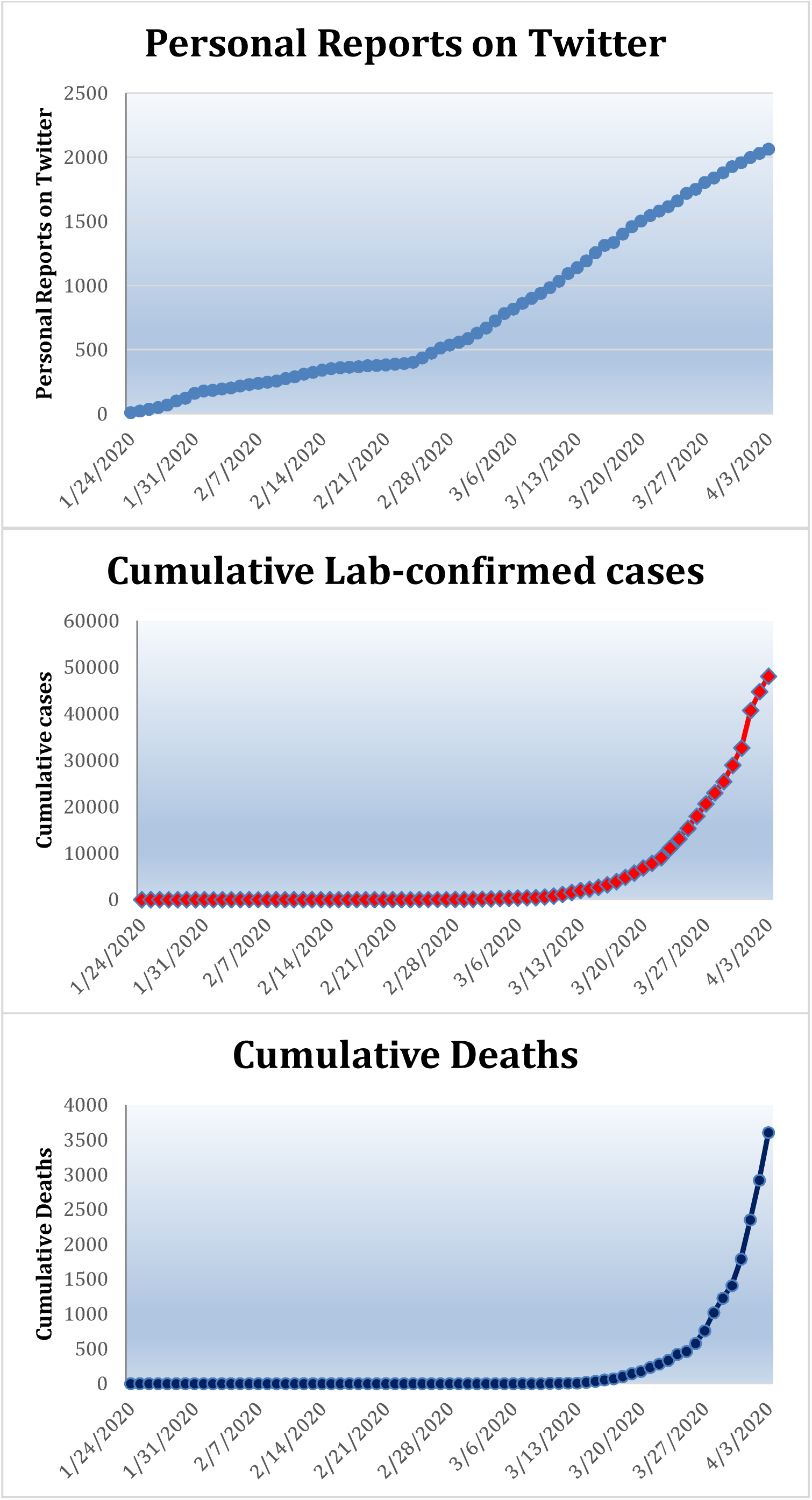
Cumulative number of users posting “probable” or “possible” tweets, cumulative lab-confirmed cases and cumulative deaths by report date, January 23, 2020 to April 6, 2020

**Figure 3:**
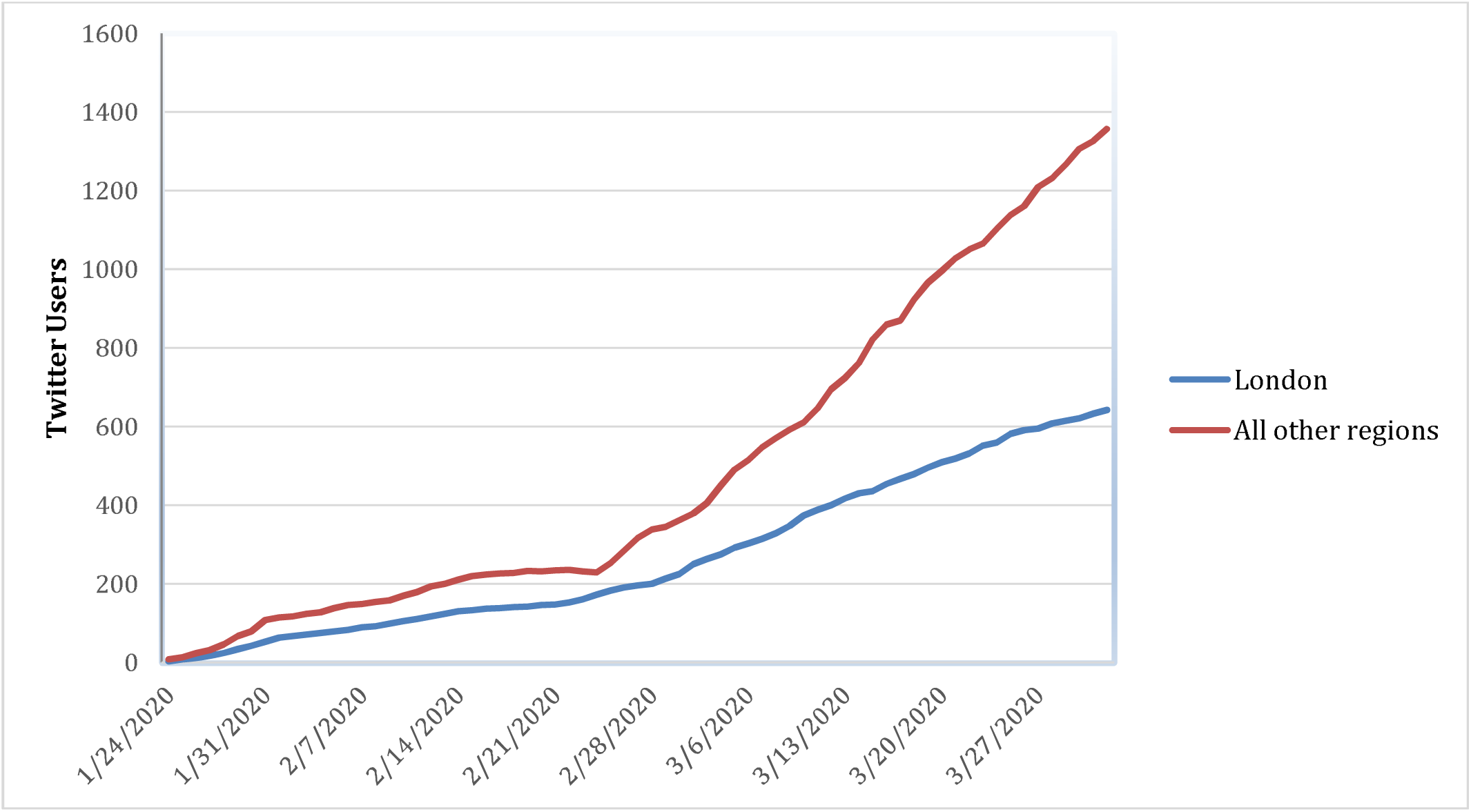
Cumulative number of users posting “probable” or “possible” tweets, and lab-confirmed cases with a London or other region in England by report date, January 23, 2020 to April 6, 2020

## Discussion

In Figure 1, there is overall agreement in the proportion of users in Twitter by region and those from UK government statistics of confirmed cases. These personal reports on Twitter began to increase sharply around the beginning of March, as shown in Figure 2, but not until the middle/end of March for UK government confirmed cases or deaths. There is steeper curve for personal reports on Twitter outside London than within London in Figure 3.

This is similar to the findings for reports in the United States. Indeed similarly to the United States, we have detected “probable” or “possible” tweets that were posted before the region’s first confirmed case. This raises more question than answers but is an interesting finding given that the first confirmed cases and deaths are now thought to be earlier than first recorded.

The potential for using Twitter data has not yet been realized and could include analysis of the temporal and geographic range and types of symptoms in different populations and geographical locations identified may also lend themselves to more precise data for analysis.

Thus, real-time, user-generated Twitter data could help provide an early indication of the spread of COVID-19, not just in the US but also in the UK.

## Data Availability

The annotated data that was used to train the classifier for the evaluation in this study is available as a supplemental file with this article. Tweets annotated as “other,” “probable,” and “possible” are labeled as “0,” “1,” and “2,” respectively. To download the tweets, a Python script is available at https://bitbucket.org/pennhlp/twitter_data_download/src/master/.

## Conflict of Interest Disclosures

None reported.

## Funding/Support

This work was supported by the National Institutes of Health (NIH) National Library of Medicine (NLM) grant R01LM011176 and National Institute of Allergy and Infectious Diseases (NIAID) grant R01AI117011.

## Disclaimer

The content is solely the responsibility of the authors and does not necessarily represent the views of the NIH, NLM, or NIAID.

## Additional Contributions

Alexis Upshur, BS, a full-time, professional annotator in the Department of Biostatistics, Epidemiology, and Informatics at the University of Pennsylvania, contributed to annotating the Twitter data, and did not receive additional compensation for her work beyond her salary.

## Additional Information

The code for downloading the tweets in the annotated training data is available at https://bitbucket.org/pennhlp/twitter_data_download/src/master/.

## Notes

### Competing Interest Statement

The authors have declared no competing interest.

